# Higher Rate of SBP Recurrence with Secondary SBP Prophylaxis Compared to No Prophylaxis in Two National Cirrhosis Cohorts

**DOI:** 10.1101/2024.06.17.24309043

**Authors:** Scott Silvey, Nilang R Patel, Stephanie Y. Tsai, Mahum Nadeem, Richard K Sterling, John D Markley, Evan French, Jacqueline G O’Leary, Jasmohan S Bajaj

## Abstract

**Objective:** Changes in bacteriology of spontaneous bacterial peritonitis (SBP) has been documented. Reappraisal of primary SBP prophylaxis showed an increased rate of resistance in patients on primary prophylaxis with resultant discontinuation of this prophylaxis throughout the VA. We aimed to re-evaluate the risk-benefit ratio of secondary SBP prophylaxis (SecSBPPr).

**Design:** Using validated ICD 9/10 codes, we utilized the VA Corporate Data Warehouse and the Non-VA National TriNetX database to identify patients in two different large US systems who survived their first SBP diagnosis (with confirmatory chart review from two VA centers) between 2009-2019. We evaluated the prevalence of SecSBPPr and compared outcomes between those started on SecSBPPr versus not.

**Results:** We identified 4673 Veterans who survived their index SBP episode; 54.3% of whom were prescribed SecSBPPr. Multivariable analysis showed higher SBP recurrence risk in those on vs. off SecSBPPr (HR-1.63, p<0.001). This was accompanied by higher fluroquinolone-resistance risk in patients on SecSBPPr (OR=4.32, *p*=0.03). In TriNetX we identified 6708 patients who survived their index SBP episode; 48.6% were on SecSBPPr. Multivariable analysis similarly showed SecSBPPr increased the risk of SBP recurrence (HR-1.68, *p*<0.001). Both groups showed higher SBP recurrence trends over time in SecSBPPr patients.

**Conclusion:** In two national data sets of >11,000 patients with SBP we found that SecSBPPr was prescribed in roughly half of patients. When initiated, SecSBPPr, compared to no prophylaxis after SBP, increased the risk of SBP recurrence in multivariable analysis by 63-68%, and this trend worsened over time. SecSBPPr should be reconsidered in cirrhosis.

• **What is already known on this topic –**

➢ Secondary prophylaxis to prevent recurrence of spontaneous bacterial peritonitis (SBP) has been recommended in several guidelines,
➢ Changing demographics and bacteriology could impact the effectiveness of secondary SBP prophylaxis, but a national perspective is needed.
➢ In a national Veterans cohort, primary SBP prophylaxis was associated with worse outcomes due to antibiotic resistance, which led to the VA discouraging this practice system-wide. However, the data regarding SBP prophylaxis is unclear.
• **What this study adds –**

➢ Almost 50% of patients with cirrhosis with SBP across 2 large US-based National cohorts (Veterans and TriNetX) evaluated from 2009-2019 were not initiated on secondary SBP prophylaxis, which gave us an opportunity to analyze the effectiveness over time in preventing recurrence.
➢ In >11,000 patients regardless of Veterans or non-Veterans, the use of secondary SBP prophylaxis worsened the rate of SBP recurrence without changes in mortality compared to those who were not on it.
➢ The SBP recurrence rate with secondary SBP prophylaxis worsened as time progressed in both cohorts and was associated with worsening antibiotic resistance.
• **How this study might affect research, practice, or policy –**

➢ The lack of improvement and higher SBP recurrence in patients on secondary SBP prophylaxis spanning two complementary cohorts should lead policymakers and antimicrobial stewardship professionals to re-evaluate the utility of this practice.
➢ Focusing on increasing ascites fluid culture to select patients who could benefit from secondary SBP prophylaxis may be necessary.

## Introduction

With advancing cirrhosis and development of ascites, there is a risk of spontaneous bacterial peritonitis (SBP)[1]. Delays in or absence of SBP treatment is associated with acute kidney injury, organ failure(s), acute-on-chronic liver failure and death, and once SBP occurs, there is a high rate of recurrence[2, 3, 4]. The microbiome in patients with cirrhosis changes as liver disease progresses, with etiology of liver disease, and with medications given. Patients with decompensated cirrhosis have high rates of bacterial, fungal, and viral dysbiosis, which is worsened by antibiotic use, such as SBP prophylaxis, and leads to further enrichment of antibiotic resistance genes[5, 6, 7, 8]. The current preventive strategies for recurrence include daily antibiotic use, usually with fluoroquinolones or trimethoprim-sulfomethoxazole (TMP-SMX)[1]. However, with the increasing prevalence of antibiotic resistance, as well as the shift in SBP causative organisms from gram-negative to gram-positive organisms, the real-world efficacy of SBP prophylaxis needs to be re-examined[9, 10]. This re-evaluation is particularly relevant now, since recent data documented the risk-benefit ratio of primary SBP prophylaxis has changed, likely secondary to the increasing prevalence of resistant organisms[11, 12, 13].

Therefore, it is time to re-evaluate the risks and benefits of secondary SBP prophylaxis (SecSBPPr). This re-appraisal of SecSBPPr needs analysis of large nationally representative cohorts to ensure that differences in practice patterns do not dictate results. To address this gap in knowledge, two cohorts were studied: the Corporate Data Warehouse (CDW) of US Veterans and the non-Veteran TriNetX cohort. We hypothesized the rate of SBP recurrence is higher and is worsening over time in patients with cirrhosis and prior SBP who were initiated on SecSBPPr compared to those who were not initiated on SecSBPPr after an episode of SBP in two large national US cohorts.

## Methods

We used the VA Corporate Data Warehouse (CDW) with chart review from two centers to further validate the codes for SBP, and the TriNetX database from 2009-2019.

### VA CDW

#### Cohort creation

We obtained the first (index) inpatient or outpatient diagnosis of SBP between 2009-2019 using validated ICD 9/10 codes (ICD 9: 567.23, ICD 10: K65.2) in each cohort. Patients were observed starting 30 days after the SBP diagnosis for 2 years. The latest index SBP diagnosis date considered was December 1, 2017, so that each patient received a full 2-year window of follow-up time. Patients who died or received a liver transplant up to 30 days after their index SBP infection were not included in the cohort as these outcomes were likely caused by the index SBP event and not from the presence or absence of prophylactic medication. SecSBPPr was defined as continuous use (≥2 refills) of fluroquinolones or TMP-SMX up to 120 days after the index diagnosis date. We also collected additional information on demographics, admission medications, MELD-Na, platelet count (10^9^/L), albumin (g/dL), white blood cell count (10^9^/L), Charleson Comorbidity Index (CCI), and outcomes. A full description of definitions can be found in the Supplementary section. Cohort characteristics were summarized and compared between the groups: no SecSBPPr versus SecSBPPr (Table 1). Continuous variables were presented as the mean (± SD) or median (IQR), and categorical variables as counts and percentages of the total. Variables were compared between the groups using two-sample *t*-tests, Wilcoxon Rank-Sum tests, or Pearson’s Chi-Squared tests, as appropriate.

**Table 1:** All patients with SBP in the VA CDW Database.

| ***n* = 4673 patients with first SBP episode** | **Not started on Secondary Prophylaxis (*n* = 2134, 45.7%)** | **Started On Secondary Prophylaxis (*n* = 2539, 54.3%)** | ***p*-value** |
| --- | --- | --- | --- |
| **Variable** |  |  |  |
| **Labs / Demographics** |  |  |  |
| Age | 61.49 (± 9.13) | 61.55 (± 8.11) | 0.80 |
| Male Sex | 2045 (97.7%) | 2430 (96.7%) | 0.07 |
| White Race | 1533 (77.7%) | 1912 (81.1%) | **0.005** |
| Hispanic Ethnicity | 194 (9.4%) | 252 (10.3%) | 0.36 |
| Alcohol Etiology | 812 (38.1%) | 999 (39.3%) | 0.38 |
| Charlson Comorbidity Index | 5.02 (± 2.65) | 5.52 (± 2.69) | **<0.001** |
| MELD-Na | 17.87 (± 6.78) | 18.34 (± 6.24) | **0.02** |
| Platelet Count ($10^9/L$) | 134.00 (81.00-221.00) | 101.00 (66.00-170.00) | **<0.001** |
| Albumin (g/dL) | 2.72 (± 0.67) | 2.81 (± 0.65) | **<0.001** |
| White Blood Cell Count ($10^9/L$) | 6.80 (4.80-9.30) | 5.87 (4.10-8.20) | **<0.001** |
| VA Complexity, Level 1 | 1969 (92.3%) | 2428 (95.6%) | **<0.001** |
| North Atlantic Region | 424 (19.8%) | 436 (17.2%) | **0.020** |
| **Medications** |  |  |  |
| Proton Pump Inhibitors | 772 (36.2%) | 1028 (40.5%) | **0.003** |
| Statins | 200 (9.4%) | 245 (9.6%) | 0.79 |
| Lactulose | 622 (29.1%) | 948 (37.3%) | **<0.001** |
| Rifaximin | 191 (9.0%) | 364 (14.3%) | **<0.001** |
| Propranolol | 348 (16.3%) | 501 (19.7%) | **0.003** |
| Nadolol | 41 (1.9%) | 83 (3.3%) | **0.006** |
| Carvedilol | 80 (3.7%) | 107 (4.2%) | 0.46 |
| Selective Beta-Blocker | 208 (9.7%) | 245 (9.6%) | 0.95 |
| **Outcomes** |  |  |  |
| 2-Year SBP Recurrence | 293 (13.7%) | 611 (24.1%) | **<0.001** |
| 2-Year All-Cause Mortality | 1261 (59.1%) | 1564 (61.6%) | 0.08 |
| 2-Year Liver Transplant | 33 (1.5%) | 87 (3.5%) | **<0.001** |

**Table 2 :** Univariable and Multi-Variable analyses for Outcomes in VA-CDW.

| **Univariable Analysis** |  |  |
| --- | --- | --- |
| **Outcome** | **Hazard or Odds Ratio (95% CI)** | **p-value** |
| SBP recurrence | 1.82 [1.59-2.10] | **<0.001** |
| All-Cause Mortality | 1.11 [0.98-1.25] | 0.081 |
| Liver Transplant | 2.13 [1.52-3.43] | **<0.001** |
| **Multi-Variable analysis** |  |  |
| **Outcomes** | **Hazard or Odds Ratio (HR/OR) (95% CI)** | **p-value** |
| **SBP Recurrence** |  |  |
| Secondary Prophylaxis | 1.63 [1.40-1.91] | **<0.001** |
| Platelet count | 0.99 [0.99-1.00] | **<0.001** |
| MELD-Na | 1.03 [1.02-1.04] | **<0.001** |
| **All-Cause Mortality** |  |  |
| Secondary Prophylaxis | 0.93 [0.81-1.06] | 0.28 |
| Platelet count | 1.00 [0.99-1.00] | **<0.001** |
| Proton Pump Inhibitors | 1.21 [1.05-1.40] | **0.009** |
| Lactulose | 1.20 [1.03-1.40] | **0.02** |
| North Atlantic Region | 0.82 [0.69-0.98] | **0.03** |
| Charlson Comorbidity Index | 1.20 [1.17-1.24] | **<0.001** |
| WBC count | 1.02 [1.00-1.04] | **0.03** |
| MELD-Na | 1.06 [1.05-1.07] | **<0.001** |
| Albumin | 0.62 [0.56-0.69] | **<0.001** |
| **Liver Transplant** |  |  |
| Secondary Prophylaxis | 1.85 [1.20-2.94] | **0.007** |
| Platelets | 1.00 [0.99-1.00] | **0.03** |
| North Atlantic Region | 1.70 [1.06-2.65] | **0.02** |
| Rifaximin | 1.88 [1.17-2.96] | **0.007** |
| MELD-Na | 1.06 [1.03-1.09] | **<0.001** |
| WBC count | 0.89 [0.81-0.97] | **0.008** |

Death/transplant were odds ratios and time to recurrence was hazards ratio

<u>Analysis of Outcomes</u>: The primary outcome of interest was the time (in days) to SBP recurrence during the 2-year follow-up period that was > 30 days after the index episode. We included this one-month buffer period between the index date and the beginning of the follow-up period to ensure that the second SBP episode was truly a new instance of the disease and the outcomes related to new events. We used Cox Proportional Hazards models to examine this outcome, and patients who did not experience recurrence during follow-up were censored, as were patients who died or received liver transplant prior to recurrence. All-cause mortality and liver transplant rates were examined as secondary outcomes; these were examined using logistic regression models.

#### Change over time

For all models, we also adjusted for covariates that were significantly different between the no-SecSBPPr versus SecSBPPr groups. Finally, in the SBP recurrence model, we hypothesized that the potential effect of secondary SBPPr on SBP recurrence would become worse over time. Thus, an additional interaction between the time from index diagnosis (defined numerically as the number of days after Jan 1, 2009 of the patient’s index date, divided by 365) and secondary prophylaxis was tested.

#### Antibiotic Resistance

We examined the proportion of fluoroquinolone-resistant infections among those on fluroquinolone-specific SecSBPPr (vs. no SecSBPPr), as well as the proportion of TMX-SMP-resistant infections among those on TMP-SMX SecSBPPr (vs. no SecSBPPr). Antibiotic resistance data was collected within the date of recurrence ±14 days to capture all potential susceptibility results within the infection date. Logistic regression models were used to examine associations, and further multivariable modeling was performed, again adjusting for all covariates that were significantly different between the secondary prophylaxis groups.

RStudio version 4.3.1 was used for all statistical analysis. All hypothesis tests were two-sided with statistical significance considered *p* < 0.05.

Validation of results in two VA Centers: Charts from the patients included in the CDW from the Richmond and Dallas VA were reviewed to confirm the presence of SBP and whether fluoroquinolone or TMP-SMX use was indeed started for SecSBPPr.

### Validation of Results in a TriNetX National Non-Veteran Cohort

TriNetX is a national database of insured, non-VA patients (trinetx.com) sourced from health care organizations (HCOs) participating in the TriNetX Research Network. These HCOs are usually large academic medical institutions with both inpatient and outpatient facilities within the US. TriNetX contains information regarding patient demographics, diagnoses and procedures through ICD or CPT codes, laboratory values, prescription data, transplant, and death records. This cohort was assembled using similar methods as described above, but with some differences. First, some covariates were not available (or very sparse) in TriNetX, including hospital complexity, region, INR labs, CCI, and information on antibiotic resistance. Because INR was not available to calculate MELD-Na score, we used the MELD-XI score instead, which excludes INR. We also adjusted for individual laboratory results (if statistically significantly different between the SecSBPPr groups)[14]. Secondly, the death records on TriNetX only provide the month of death, rather than an exact date; to counteract this, we used the middle (15^th^) of each month as the day of for death-based analyses. Finally, prescription refill data was not available, so we only considered a single instance of Ciprofloxacin/TMP-SMX at the time of index diagnosis (up to 120-days post-index episode) to be indicative of SecSBPPr. Identical statistical analyses were performed on TriNetX as VA CDW cohort, and the results were compared.

IRB approval was obtained from the Richmond and Dallas VA (for the VA records) and VCU (for TriNetX) Institutional Review Boards before the study was initiated. The IRB approvals included waiver of individual informed consent.

Patient and public involvement in research: There was no involvement of patients or public in this research design.

## Results

### VA-CDW Cohort Creation and Description

From 2009-2019, we identified 4673 patients who survived their index SBP episode, with 2539 (54.3%) started on secondary SBP prophylaxis after the index episode. Among these, 2144 (84.4%) were on fluoroquinolones (Ciprofloxacin: 1985, 92.6%, Other: 159, 7.4%) and 395 (14.6%) were on TMP-SMX. Cohort characteristics can be found in Table 1. Patients who were put on SecSBPPr were significantly more likely to be on lactulose (37.3% vs. 29.1%, *p*<0.001), rifaximin (14.3% vs. 9.0%, *p*<0.0001), propranolol (19.7% vs. 16.3%, *p*<0.001), nadolol (3.3% vs. 1.9%, *p*=0.006), and proton pump inhibitors (PPI) (40.5% vs. 36.2%, *p*=0.003). These patients also had higher mean CCI (5.52 vs. 5.02, *p<*0.001), serum albumin (2.81 vs. 2.72, *p*<0.001), and MELD-Na scores (18.34 vs. 17.87, *p*=0.017); lower median WBC counts (5.87 vs. 6.80, *p*<0.001) and median platelet counts (101.00 vs. 134.00, *p*<0.001); were more likely to be White (81.1% vs. 77.7%, *p*=0.005), in VA complexity level 1 centers (95.6% vs. 92.3%, *p*<0.001), and less likely to be admitted in the North Atlantic VA Region (17.2% vs. 19.9%, *p*=0.020).

#### VA-CDW Multivariable Analysis

Among the 4673 veterans who survived their index SBP episode, 904 (19.3%) had a second SBP episode within the 2-year follow-up period. The crude rates of recurrence (24.1% vs. 13.7%, p<0.001) and liver transplant rate (3.5% vs. 1.5%, p<0.001) were higher in those on vs. off SecSBPPr (Table 1). On univariable analysis, the rates of SBP recurrence in the SecSBPPr group were significantly higher (Figure 1, HR: 1.82, 95% CI: [1.59-2.10], *p*<0.001), as were the odds of liver transplant (OR: 2.13, 95% CI: [1.52-3.43], *p*<0.001). The odds of all-cause mortality in the SecSBPPr group were numerically higher but not statistically significant (OR: 1.11, 95% CI: [0.98-1.25], *p*=0.081).

**Figure 1:**
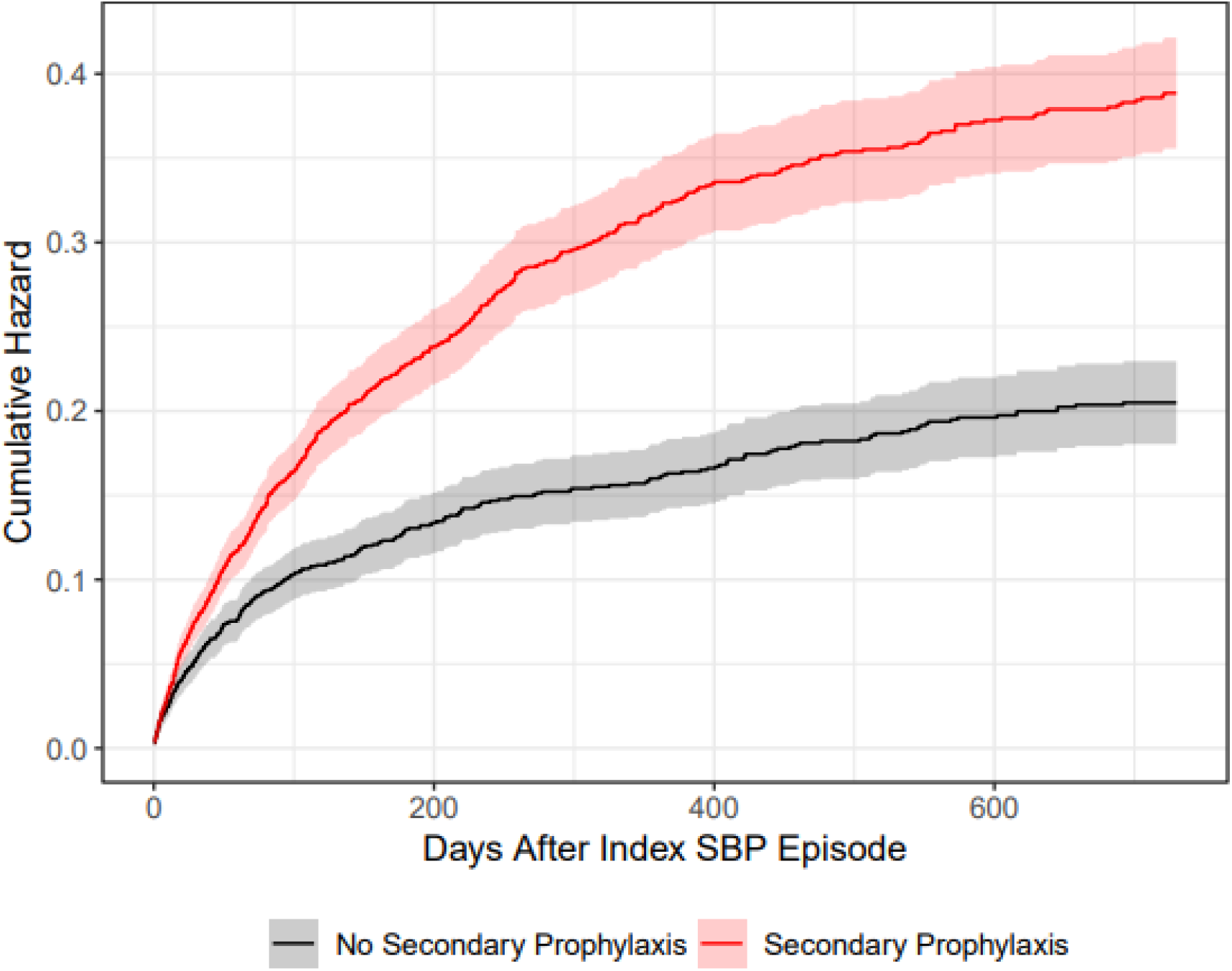
Cumulative Incidences for SBP recurrence in Veterans Affairs Corporate Data Warehouse. Data are presented as cumulative hazards for SBP recurrence with solid curves and 95% CI shading. Patients on secondary prophylaxis (red) had an adjusted hazard ratio of 1.62 (95% CI: 1.40-1.91, p<0.0001) for SBP recurrence versus those who were not on secondary prophylaxis (gray) adjusted for significantly different clinical covariates.

After adjusting for all covariates that were significantly different between the treatment groups, the rates of SBP recurrence remained statistically significantly higher in patients on SecSBPPr (*p*<0.001, Adj. HR: 1.63, 95% CI: [1.40-1.91]). Other variables associated with a higher risk of a second SBP episode were lower platelet counts (*p*<0.001) and higher MELD-Na score (*p*<0.001).

#### Analysis of Time period

When the interaction between continuous time and SecSBPPr were added to the adjusted Cox Proportional Hazards model for SBP recurrence, we found that for patients receiving SecSBPPr were 1.07 times more likely to have a second SBP episode for every additional year post-2009 (95% CI: [1.01 – 1.13], *p*=0.026). Visual evidence of this trend can be seen in Figure 2.

**Figure 2:**
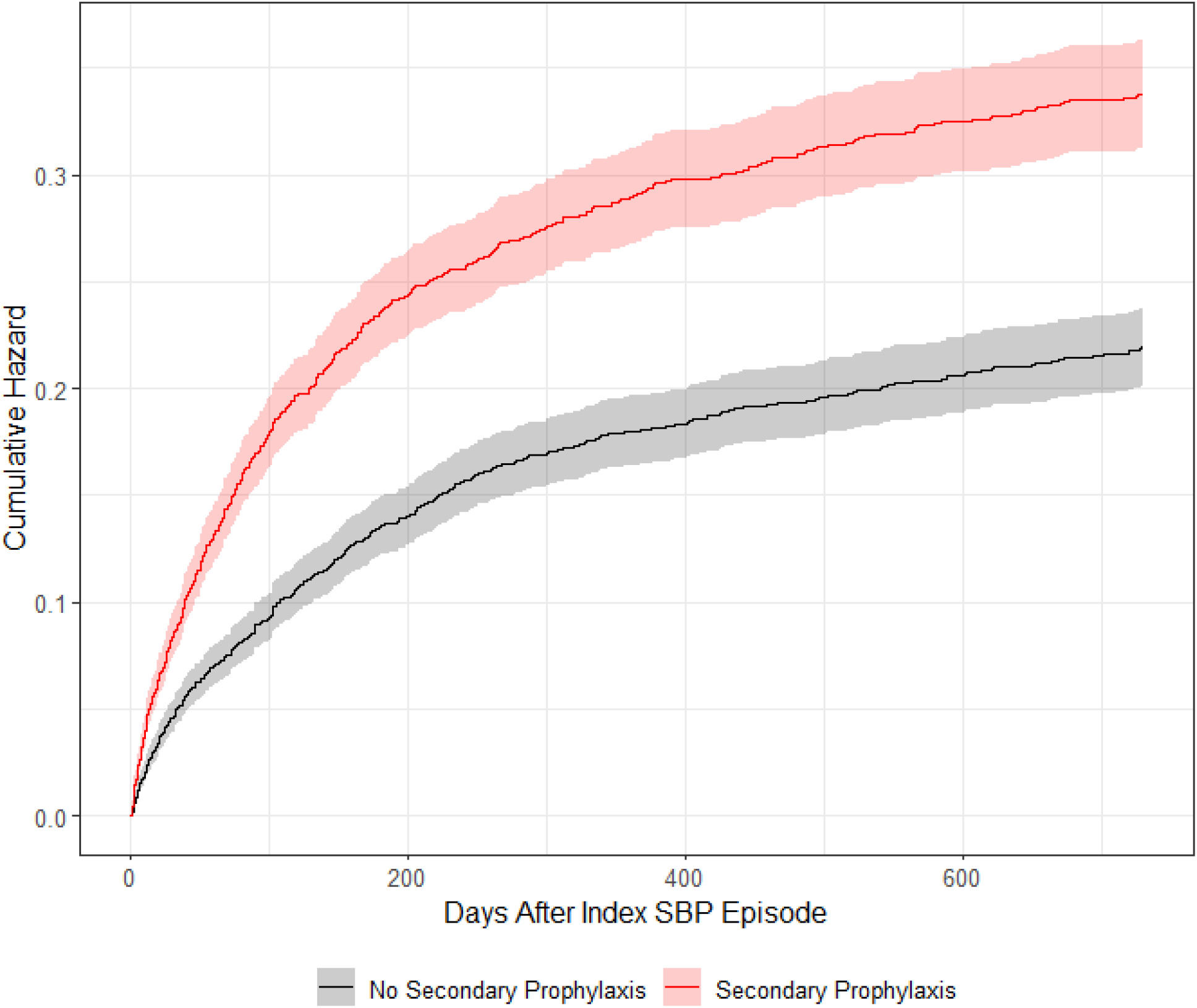
Cumulative Incidences for SBP Recurrence in the TriNetX Cohort. Data are presented as cumulative hazards for SBP recurrence with solid curves and 95% CI shading. Patients on secondary prophylaxis (red) had an adjusted hazard ratio of 1.68 (95% CI: 1.33-1.80, p<0.0001) for SBP recurrence versus those who were not on secondary prophylaxis (gray) adjusted for significantly different clinical covariates.

#### Other outcomes

For all-cause mortality, adjusting for covariates did not change the results; death rates were not altered by the presence or absence of SecSBPPr on multi-variable analysis (Table 2, *p*=0.28). Other variables associated with higher rates of death were PPI use (*p*=0.008), lactulose use (*p*=0.021), regions other than North Atlantic (*p*=0.025), higher MELD-Na (*p*<0.001), higher CCI (*p*<0.001), lower platelet counts (*p*<0.001), higher WBC (*p*<0.001), and lower albumin (*p*<0.001).

For liver transplant, after adjusting for covariates, the positive association between secondary prophylaxis and this outcome remained consistent (adj. OR: 1.85, 95% CI: [1.20-2.94], *p*=0.007). Other variables associated with higher transplant rates were North Atlantic Region (*p*=0.023), rifaximin use (*p*=0.007), higher MELD-Na (*p*<0.001), lower platelets (*p*=0.032) and lower WBC counts (*p*=0.008). For all adjusted models, specific hazard/odds ratios and 95% confidence intervals can be found in Table 2.

#### Antibiotic Resistance

Only 100 (2.2%) patients were culture-positive with antibiotic resistance data available. Of these patients, all had sensitivity data for fluroquinolones, and 16 for TMP-SMX sensitivity. Within those on fluroquinolone SecSBPPr, 67.9% (38/56) had evidence of fluroquinolone-resistant isolates versus 45.5% in those not on SecSBPPr (OR: 2.53, 95% CI: [1.13-5.82], *p*=0.026). Multivariable analysis of fluroquinolone resistance documented SecSBPPr to be statistically significantly associated with this outcome after adjusting for all covariates previously discussed (adj. OR: 4.32, 95% CI: [1.36-15.83], *p*=0.018). There was only one patient on TMP-SMX SecSBPPr within the subset with sensitivity data; this patient did have a TMP-SMX-resistant infection (N=1, 100.0%) versus 46.6% (7 of 15) resistance in those not on TMP-SMX SecSBPPr, but this sample size was insufficient for further statistical analysis.

#### Chart review

The Dallas VA cohort had 90 patients and Richmond VA had 45 patients that were included in the CDW with a computer diagnosis of SBP. Within the Dallas VA, the mean age was 61.7±7.6 years and all patients except 2 were male. Documented SBP was seen in 78% (70/90) of patients, 7% (6/90) of patients were treated empirically for undocumented SBP, and the other patients had other types of infections during their admission. Of these SBP patients, 67% were started on SecSBPPr (n= 41 on a fluroquinolone, n=3 on TMP-SMX and n=3 on cefpodoxime).

In the Richmond VA cohort, the mean age was 60.4±9.6 years, and only one patient was a woman. 80% (36 of 45) had SBP; the rest were unclear (n=5) or had secondary SBP due to umbilical hernia incarceration or hepatic abscesses (n=4). Of the 36 SBP patients, 26 were started on SecSBPPr (n=21 on fluoroquinolones and n=5 on TMP-SMX). Ascites fluid organisms were isolated in 36% (n=13) of the SBP patients (9 gram-negative, 3 gram-positive and one fungus).

### TriNetX cohort

This cohort contained 6708 patients with SBP (age 56.42 ± 11.23, 64.2% male, 48.6% on SecSBPPr). Patients on SecSBPPr had similar trends in cohort characteristics; namely greater severity of cirrhosis and higher rates of admission medications (Rifaximin, Lactulose, PPI Table 3). SecSBPPr patients were also more likely to white, of non-Hispanic ethnicity, and have an alcohol-related etiology of cirrhosis. On univariable analysis (Table 4), SecSBPPr patients again had higher rates of 2-year SBP recurrence (HR: 1.61 [1.44 – 1.80], *p*<0.001), LT (OR: 2.86 [2.40 – 3.41], *p*<0.001), and death (OR: 1.16 [1.05 – 1.29], *p*=0.003).

**Table 3:** All patients with SBP in the TriNetX Database.

| ***n* = 6708 patients with first SBP episode** | **Not started on Secondary Prophylaxis (*n* = 3447, 51.4%)** | **Started On Secondary Prophylaxis (*n* = 3261, 48.6%)** | ***p*-value** |
| --- | --- | --- | --- |
| **Variable** |  |  |  |
| **Labs / Demographics** |  |  |  |
| Age | 56.66 (± 11.47) | 56.17 (± 10.96) | 0.07 |
| Male Sex | 2191 (63.6%) | 2116 (64.9%) | 0.27 |
| White Race | 2260 (76.7%) | 2356 (79.7%) | **0.005** |
| Hispanic Ethnicity | 598 (22.7%) | 495 (19.2%) | **0.002** |
| Alcohol Etiology | 1739 (50.4%) | 1829 (56.1%) | **<0.001** |
| MELD-XI Score | 16.84 (± 6.34) | 16.93 (± 5.79) | 0.62 |
| Bilirubin (g/dL) | 1.8 (0.9-3.8) | 2.4 (1.2-4.6) | **<0.001** |
| Creatinine (mg/dL) | 1.00 (0.72-1.60) | 0.93 (0.70-1.31) | **<0.001** |
| Sodium (mEq/L) | 135.03 (± 5.39) | 134.73 (± 5.38) | **0.04** |
| Albumin (g/dL) | 2.83 (± 0.70) | 2.77 (± 0.66) | **0.006** |
| White Blood Cell Count ($10^9$/L) | 6.50 (4.40-9.61) | 6.20 (4.20-9.00) | **0.01** |
| Platelet Count ($10^9$/L) | 116.00 (73.00-187.00) | 100.00 (65.00-159.00) | **<0.001** |
| **Medications** |  |  |  |
| Proton Pump Inhibitor | 1331 (38.6%) | 1734 (53.2%) | **<0.001** |
| Statins | 262 (7.6%) | 263 (8.1%) | 0.51 |
| Lactulose | 1154 (33.5%) | 1605 (49.2%) | **<0.001** |
| Rifaximin | 600 (17.4%) | 937 (28.7%) | **<0.001** |
| Propranolol | 281 (8.2%) | 389 (11.9%) | **<0.001** |
| Nadolol | 201 (5.8%) | 376 (11.5%) | **<0.001** |
| Carvedilol | 145 (4.2%) | 128 (3.9%) | 0.60 |
| Selective Beta-Blocker | 299 (8.7%) | 325 (10.0%) | 0.08 |
| **Outcomes** |  |  |  |
| 2-Year SBP Recurrence | 551 (16.0%) | 734 (22.5%) | **<0.001** |
| 2-Year All-Cause Mortality | 1157 (33.6%) | 1207 (37.0%) | **0.003** |
| 2-Year Liver Transplant | 191 (5.5%) | 468 (14.4%) | **<0.001** |

**Table 4:**
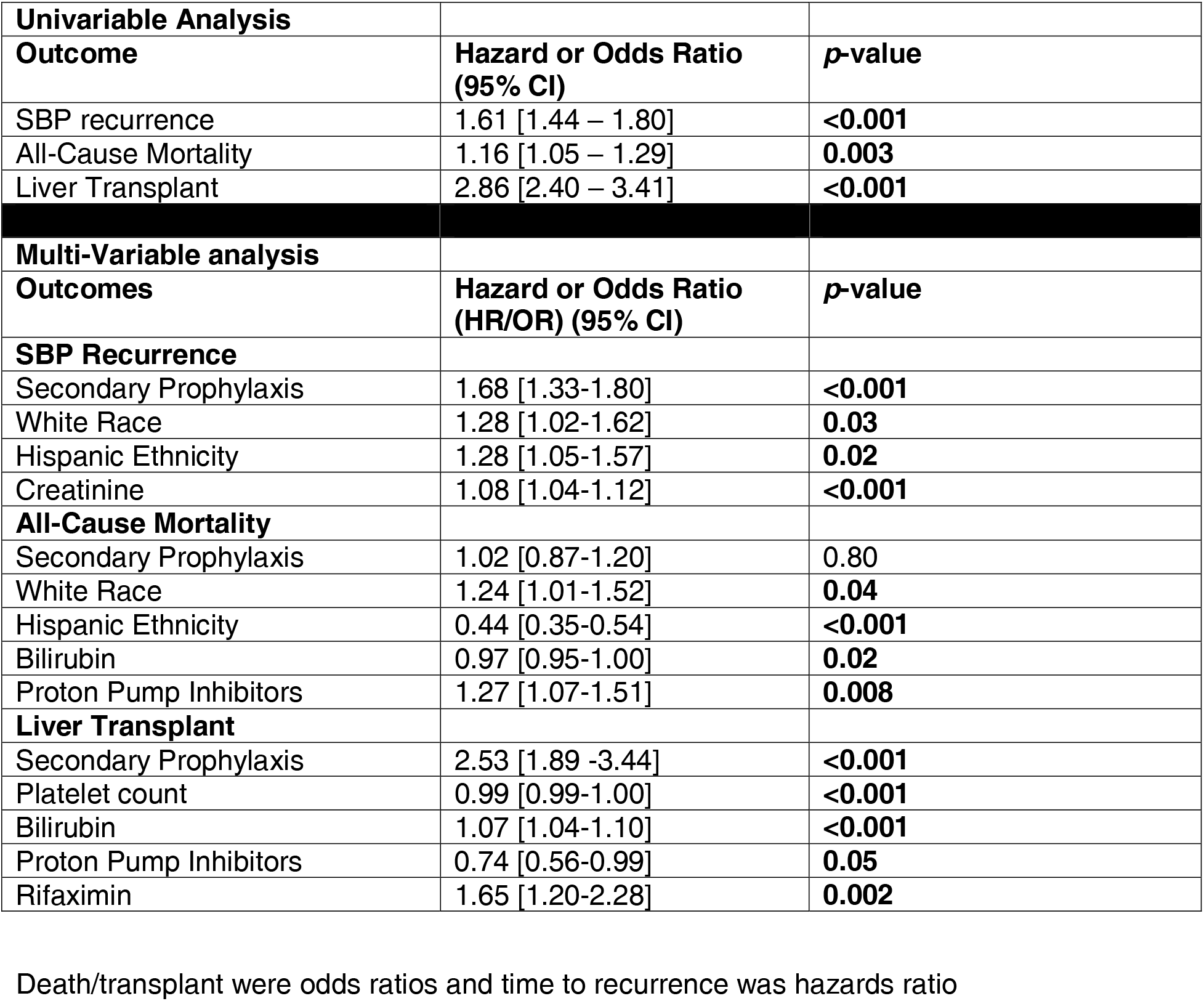
Univariable and Multi-Variable analyses for Outcomes in TriNetX.

| **Univariable Analysis** |  |  |
| --- | --- | --- |
| **Outcome** | **Hazard or Odds Ratio (95% CI)** | **p-value** |
| SBP recurrence | 1.61 [1.44 – 1.80] | **<0.001** |
| All-Cause Mortality | 1.16 [1.05 – 1.29] | **0.003** |
| Liver Transplant | 2.86 [2.40 – 3.41] | **<0.001** |
| **Multi-Variable analysis** |  |  |
| **Outcomes** | **Hazard or Odds Ratio (HR/OR) (95% CI)** | **p-value** |
| **SBP Recurrence** |  |  |
| Secondary Prophylaxis | 1.68 [1.33-1.80] | **<0.001** |
| White Race | 1.28 [1.02-1.62] | **0.03** |
| Hispanic Ethnicity | 1.28 [1.05-1.57] | **0.02** |
| Creatinine | 1.08 [1.04-1.12] | **<0.001** |
| **All-Cause Mortality** |  |  |
| Secondary Prophylaxis | 1.02 [0.87-1.20] | 0.80 |
| White Race | 1.24 [1.01-1.52] | **0.04** |
| Hispanic Ethnicity | 0.44 [0.35-0.54] | **<0.001** |
| Bilirubin | 0.97 [0.95-1.00] | **0.02** |
| Proton Pump Inhibitors | 1.27 [1.07-1.51] | **0.008** |
| **Liver Transplant** |  |  |
| Secondary Prophylaxis | 2.53 [1.89 -3.44] | **<0.001** |
| Platelet count | 0.99 [0.99-1.00] | **<0.001** |
| Bilirubin | 1.07 [1.04-1.10] | **<0.001** |
| Proton Pump Inhibitors | 0.74 [0.56-0.99] | **0.05** |
| Rifaximin | 1.65 [1.20-2.28] | **0.002** |

Death/transplant were odds ratios and time to recurrence was hazards ratio

#### Multivariable adjustment

After covariate adjustment, the rate of 2-yr SBP recurrence remained significantly higher in SecSBPPr (HR: 1.68 [1.33-1.80], *p*<0.001), as did the odds of liver transplant (OR: 2.53 [1.89 -3.44], *p*<0.001). Mortality was no longer statistically significant (*p*=0.795).

Change over time: When the interaction between continuous time and SecSBPPr were added to the adjusted Cox Proportional Hazards model for SBP recurrence, we found that for patients receiving SecSBPPr were 1.11 times more likely to have a recurrent SBP episode for every additional year post-2009 (95% CI: [1.03 – 1.21], *p*=0.011).

## Discussion

In two large US-based cohorts of patients, both veterans and non-veterans with cirrhosis and SBP, study data demonstrated that those who were initiated on SecSBPPr after the index case of SBP had a higher rate of SBP recurrence compared to those that did not receive prophylaxis. Moreover, this higher rate of SBP recurrence increased in those who received SecSBPPr over time, with the highest separation being in the latter time-periods.

These two cohorts of patients demonstrated that only roughly half of patients after SBP diagnosis are placed on secondary SBP prophylaxis per AASLD guidance and EASL guidelines[1, 15]. Likely patient, provider and systems problems are at play in this relatively low rate of utilization of what is believed to be an important quality metric. Patient-based reasons may include non-adherence with follow up, low health literacy, fear of side effects, or cost of the medication. And system level problems may include but are not limited to lack of education of the importance of prophylaxis to midlevel or primary care providers, lack or records availability or communication if a patient was diagnosed at an outside hospital.

Since SBP prophylaxis is considered current standard of care, only database studies can challenge this ingrained dogma. Due to the marked changes in etiology of liver disease, access to liver transplant, microbiology resistance rates and SBP causative organisms since original studies of primary and secondary SBP prophylaxis in the 1980s and 1990s we sought to re-evaluate the risk-benefit ratio of these prophylactic therapies[16]. Our first step was a reappraisal of primary SBP prophylaxis. This undertaking first reveled that outcome of prospectively enrolled inpatients in the North American Consortium for the Study of End-stage Liver Disease admitted on primary SBP prophylaxis vs. secondary SBP prophylaxis were worse, even after propensity score matching[12]. In fact, patients on primary SBP prophylaxis had a higher rate of SIRS, need for ICU care, nosocomial SBP rate, readmission rate, and inpatient and 90-day mortality rate. We then evaluated the national VA CDW data to evaluate outcomes of patients with SBP who were on vs. off primary SBP prophylaxis. Patients taking primary SBP prophylaxis who developed SBP had a much higher rate of gram-negative resistance. European data has also documented a declining benefit of primary SBP prophylaxis with Norfloxacin no longer having a survival advantage in a randomized controlled trial[17]. Given the changing risk-benefit ratio of primary prophylaxis combined with the high rate of *E. coli* resistance across the US to ciprofloxacin the National VA Health Care System decided to no longer recommend primary SBP prophylaxis[11].

Therefore, the next step was to re-evaluate the utility of secondary SBP prophylaxis. Interestingly, those initiated on SecSBPPr had a *higher* rate of SBP recurrence compared to those not receiving SecSBPPr. Moreover, this higher SBP recurrence numerically increased in those who received secondary SBP prophylaxis over time. This database appraisal provided an opportunity to assess the impact of SecSBPPr on important clinical outcomes such as SBP recurrence, death, and liver transplant. Our major finding was that despite controlling for clinical, patient-based, and system-based factors, SecSBPPr use emerged as a significant contributor to SBP recurrence without any mortality benefit. This is striking because the initial randomized clinical trials and guidelines cite prevention of SBP recurrence as the primary reason for SecSBPPr initiation[1, 10, 16].

Reasons for this higher rate of SBP recurrence in SecSBPPr are likely related to diminishing coverage of causative organisms over time due to increased prevalence of resistance and/or change in microbiology[13, 18, 19, 20, 21]. With recent studies in patients on SecSBPPr there was a higher relative abundance of gram-positive pathobionts, increase in microbial virulence, and changes in bacteriophage linkages that could affect effectiveness of antibiotics[13, 22, 23]. In the VA-CDW database, this was found in those who had culture data available. The results are also consistent with the VA-CDW experience in primary SBP prophylaxis, which showed up to 50% fluoroquinolone resistance to *E. coli* and *K. pneumoniae* that are main targets for SBP prophylaxis[11]. Another indirect piece of evidence is the widening of the gap in SBP recurrence rate over time, with a higher rate in SecSBPPr over time. There is ample evidence of the worsening resistance profile and shift from gram-negative to gram-positive organisms in outpatients and inpatients with cirrhosis, which is consistent with this widening of the gap over time. The sparse culture results diminish the ability of practitioners to tailor SecSBPPr strategies and instead force them to use the same regimen for all patients. It is unclear from our data how often culture bottles were inoculated at the bedside to optimize culture results, since a higher rate of culture positive SBP patients could help guide which antibiotic agent(s) to use for SecSBPPr[24].

The higher rate of SBP recurrence with SecSBPPr was consistent across two different healthcare systems, which adds to the reliability of the results. Veterans included in CDW tend to be older and male, and less likely to be minorities, compared to the general US population, while TriNetX is more reflective of the US patient population[25]. Regardless of these differences, the low rate of SecSBPPr and the pattern of consequences were similar. This included higher SBP recurrence but no significant impact on overall mortality when adjusted for baseline severity of liver disease and system-based factors. While LT was higher in the SecSBPPr group, less than 5% of patients underwent this procedure, and this small number of patients, surely followed by transplant hepatologists, may simply have been more likely to have received guideline-based care[26]. Mortality on the other hand was higher in the SecSBPPr group on crude comparisons, but not on multi-variable analysis. This is likely due to factors other than SBP that affect risk of death that are not modifiable by SecSBPPr use[27]. Patients with cirrhosis are prone to several complications that affect mortality, and these findings show the limitations of focusing on prevention of one complication[28]. Regardless, as mentioned above, the primary aim of SecSBPPr is to prevent SBP recurrence, the opposite of which was seen in our results. SBP recurrence leads to more hospitalizations, interventions, and further antibiotic use, which need to be avoided.

The question that now arises is how to decrease the rate or SBP recurrence considering this data. Until non-antibiotic strategies that are not disruptive to the microbiome and immune system are available, reflexive initiation of SecSBPPr should be reconsidered potentially with a randomized controlled clinical trial[29]. Especially in areas of high baseline resistance to the major causative organisms, initiation of SecSBPPr should be guided by antimicrobial stewardship programs. In the VA, SBP primary prophylaxis is now discouraged because of the high prevalence of resistance to the same antibiotics used for SecSBPPr. In addition to potential lack of efficacy and negative impact on the microbiome, fluoroquinolones and TMP-SMX are associated with neurologic and hematologic adverse events, as well as drug-induced liver injury, and Achilles tendon injury[30, 31, 32]. These adverse events are especially poorly tolerated in decompensated patients and add to the reasons to re-evaluate the use of SecSBPPr[32, 33]. However, ultimately non-antibiotic strategies either though lowering portal pressure or modulating the microbiome are needed to decrease the primary and secondary risk of SBP[5, 34].

Our data is limited by the database restrictions, which do not allow for specific in-depth analysis. However, we have adjusted for these limitations by either performing chart review and by utilizing two very large and different cohorts with consistent results. Because this is not an RCT, it is possible that patients in our cohort that were initiated on SecSBPPr had unmeasured risk factors for additional episodes of SBP, however, we adjusted for the most associated risk factors available and the difference between the 2 groups remained. In the chart reviews, the appropriateness of SBP diagnoses and SecSBPPr initiation were largely validated. The culture data are sparse and likely reflect clinical practice (VA-CDW) and database limitations (TriNetX). We only used a 2-year window to determine pattern changes over time and only have data through 2019 due to the COVID-19 pandemic.

We conclude that in patients newly diagnosed with SBP, initiation of SecSBPPr is associated with a 63% to 68% higher rate of SBP recurrence in multivariable analysis compared to those who were not started on prophylaxis in two large multi-center cohorts. The higher rate of SBP recurrence with SecSBPPr increased over time, likely due to increasing prevalence of resistant organisms. Careful reconsideration of SecSBPPr in patients with SBP in areas of high baseline resistance to fluoroquinolones and TMP-SMX is needed.

## Supporting information

Supplementary section

## Data availability

Data are from databases that need IRB approval to access so are not publicly accessible.

