## Supplementary section for "Higher Rate of SBP Recurrence with Secondary SBP Prophylaxis Compared to No Prophylaxis in Two National Cirrhosis Cohorts"

**Supplementary : Data Dictionary**

| Variable Name | Description | ICD Codes / Definition |
| --- | --- | --- |
| Cirrhosis DX (Inclusion Criteria) | Inclusion criteria. History of cirrhosis prior to index SBP episode. | ICD-10 : I85.00, I85.01, I85.10, I85.11, K65.2, K70.11, K70.30, K70.31, K70.40, K70.41, K71.51, K71.7, K72.10, K72.11, K74.4, K74.60. K74.69, K76.6, K76.7, K76.81  ICD-9 : 456.0, 456.1, 456.20, 456.21, 571.2, 571.5, 572.2, 572.3, 572.4 |
| SBP Index Diagnosis | Inclusion criteria. SBP episode from Jan, 1, 2009 – Dec, 1, 2017 | ICD-9 : 567.23  ICD-10 : K65.2 |
| Secondary SBPPr | Evidence of (≥ 2 refills) of Fluroquinolones or Bactrim (other name : Sulfamethoxazole/Trimethoprim) (1 = yes, 0 = no) | VA : Up to 120 days post-index (≥ 2 prescriptions/refills)  TriNetX : Up to 120 days post-index (≥ 1 prescriptions/refills) |
| Age | Patient’s age at index diagnosis. | -- |
| Male Sex | Gender (1 = male, 0 = female). | -- |
| Alcoholic Cirrhosis | Alcoholic etiology of cirrhosis (1 = yes, 0 = no). | ICD-9 : 571.2, 571.1, 571.3  ICD-10 : K70.11, K70.30, K70.31, K70.40, K70.41, K70.9, K70.10 |
| North Atlantic Region | 1 = North Atlantic, 0 = Other | <https://www.va.gov/COMMUNITYCARE/providers/Community-Care-Network.asp> |
| VA Complexity Level | 1 = Level 1a/1b/1c, 0 = Other | <https://www.ncbi.nlm.nih.gov/books/NBK555777/> |
| Sodium | Patient’s most recent sodium lab value around index diagnosis. | VA : ±30 days around index date  TriNetX : 6-months pre-index up to 30 days post-index. Units : mEq/L or mmol/L |
| Creatinine | Patient’s most recent creatinine lab value around index diagnosis. | VA : ±30 days around index date  TriNetX : 6-months pre-index up to 30 days post-index. Units : mg/dL |
| Bilirubin | Patient’s most recent bilirubin lab value around index diagnosis. | VA : ±30 days around index date  TriNetX : 6-months pre-index up to 30 days post-index.. Units : mg/dL |
| INR | Patient’s most recent INR lab value around index diagnosis. | VA : ±30 days around index date  TriNetX : Was very sparse, so not considered. |
| Platelet Count | Patient’s most recent Platelet lab value around index diagnosis. | VA : ±30 days around index date  TriNetX : 6-months pre-index up to 30 days post-index. Units : 10^9^/L  Natural log-transformed. |
| Albumin | Patient’s most recent albumin lab value around index diagnosis. | VA : ±30 days around index date  TriNetX : 6-months pre-index up to 30 days post-index. Units : g/dL |
| Total WBC | Patient’s most recent Total WBC lab value around index diagnosis. | VA : ±30 days around index date  TriNetX : 6-months pre-index up to 30 days post-index. Units : 10^9^/L  Natural log-transformed. |
| **Medications** |  |  |
| PPI | Evidence of Omeprazole, Pantoprazole, Lansoprazole, Esomeprazole, or Rabeprazole prescription. | Within 90 days pre-index |
| Statins | Evidence of Statin (Atorvastatin, Fluvastatin, Lovastatin, Pitavastatin, Pravastatin, Rosuvastatin, Simvastatin) prescription. | Within 90 days pre-index |
| Lactulose | Evidence of Lactulose prescription. | Within 90 days pre-index |
| Rifaximin | Evidence of Rifaximin prescription. | Within 90 days pre-index |
| Propranolol | Evidence of Propranolol prescription. | Within 90 days pre-index |
| Nadolol | Evidence of Nadolol prescription. | Within 90 days pre-index |
| Carvedilol | Evidence of Carvedilol prescription. | Within 90 days pre-index |
| Selective Beta-Blocker | Evidence of Atenolol, Metoprolol, Betaxolol, Bisoprolol, Acebutolol, Nebivolol, or Pindolol prescription. | Within 90 days pre-index |
| **Outcomes** |  |  |
| 2-Year SBP Recurrence | Number of days post-index that the patient had a second episode of SBP, up to 760 days post-index (2 years + 30 day buffer). | ICD-9 : 567.23  ICD-10 : K65.2 |
| 2-Year All-Cause Mortality | Number of days post-index that the patient died, up to 760 days post-index (2 years + 30 day buffer). | VA : Vital Status Mini Table.  TriNetX : Death records. |
| 2-Year Liver Transplant | Number of days post-index that the patient received liver transplant, up to 760 days post-index (2 years + 30 day buffer). | ICD-10 : Z48.23, Z94.4, T86.49, T86.40, T86.43  ICD-9 : V42.7, V58.44 |
